# Occupational challenges of health care workers during the COVID-19 pandemic. A qualitative study

**DOI:** 10.1101/2021.05.11.21257030

**Authors:** Maren Jeleff, Marianna Traugott, Elena Jirovsky-Platter, Galateja Jordakieva, Ruth Kutalek

## Abstract

**Background:** The COVID-19 pandemic has placed a significant burden on health care systems worldwide with health care workers (HCWs) delivering care under unprecedented conditions. This study deals with HCWs’ physical, mental, emotional and professional challenges of working during the COVID-19 pandemic and seeks to understand structural determinants of those challenges.

**Methods:** We carried out an exploratory qualitative study in public and private hospitals in Vienna, Austria. HCWs such as medical doctors, qualified nursing staff, nurse assistants, technical and cleaning staff in direct and indirect contact with COVID-19 patients were included. Collected data was analyzed using content analysis.

**Findings:** We conducted 30 semi-structured interviews in person and per phone from June 2020 to January 2021. Three overall themes resulted as relevant: challenges due to lack of preparedness, structural conditions, and physical and mental health of HCWs. Lack of preparedness included missing or delayed infection prevention and control (IPC) guidelines, shortages of personal protective equipment (PPE) combined with structural conditions such as staff shortages and overworked personnel. Physical and mental strains resulted from being overworked and working permanently on alert. Further, working in PPE, facing medical uncertainties and the critical conditions of patients were challenging factors. HCWs lacked recognition on multiple levels and dealt with social stigma and avoidance behavior of colleagues, especially in the beginning of the pandemic.

**Interpretation:** To mitigate HCWs’ occupational health risks and staff turnover, we propose the following context-specific recommendations: Required medical personnel in care of COVID-19 patients, especially nursing staff, should be carefully planned and increased to avert chronic work overload. Intensive training and education in palliative care, as well as in IPC for all HCWs is important. Providing supportive supervision is as essential as appropriate recognition by higher level management and the public.

**Funding:** This article has received funding from The Vienna Science and Technology Fund (WWTF) COVID-19 Rapid Response Funding 2020. The funders did not play a role in the decision to publish the article.

## 1. Introduction

From early 2020, health care systems have been challenged worldwide due to the COVID-19 pandemic. Multiple factors such as rapid spread and limited treatment options for a formerly unknown disease, the quantity of contagious patients and prolonged duration of the pandemic pose a significant burden on health care systems. HCWs are considered a vulnerable group themselves mainly through continuous exposure while caring for patients and lack of PPE.^1^

Particularly in the beginning of the pandemic, HCWs were applauded for and heroized by the public in many countries. However, many HCWs do not identify themselves as heroes but are overworked and bear the physical and mental burden of their commitment.^2^ Initially driven by enthusiasm and optimism, most feel exhausted due to the prolonged pandemic response.^3^ HCWs have to deal with the physical and mental burden of working extensively in a highly demanding situation, struck by fear of infecting family members as well as social stigma.^4, 5^ A recently published meta-analysis found female HCWs to be especially affected by anxiety and depression and a higher prevalence of these disorders in nurses than doctors.^6^

Due to the enormous pressure globally, especially nurses are quitting their jobs as stated by the International Council of Nurses.^7^ The situation is exacerbated by structural shortages of qualified nursing staff, a problem that pre-existed the pandemic with a bottleneck of 6 million nurses worldwide.^8^

Austria is an illustrative example of how COVID-19 is posing hardship to a relatively advanced and well-equipped health care system. Austria uses the Bismarck model, with health expenditures mainly being paid from taxes and mandatory social security contributions. A fundamental feature is the comprehensive health insurance coverage (99,9%) and thus accessibility to good quality care.^9-11^ In 2017 Austria was amongst the countries with the highest number of hospital beds (7.4 beds per 1000 people), which is an indicator for available resources regarding inpatient services.^10^

Nevertheless, COVID-19 overwhelmed Austria’s health care system and to date the capacity of intensive care unit (ICU) beds is reaching its limits.^12^ Moreover, Austrian hospitals were little prepared for the pandemic, with the most prominent example of temporary shortages in PPE.^13, 14^ Meanwhile the crisis also shows how the pre-existing lack of qualified personnel, especially nursing staff, has serious effects in this emergency situation.

HCWs health, well-being and safety is paramount to a functioning health care system and to ensuring patient safety.^4^ Consequently, it is necessary to mitigate risks on multiple levels – especially staff turnover and mental health risks.^6, 15^

In this study we therefore aim to address structural determinants and HCWs’ physical, mental, emotional and professional challenges of working during the COVID-19 pandemic. Based on our results, we propose context-specific recommendations.

## 2. Methods

We conducted an exploratory qualitative study with semi-structured interviews to gain insights into HCWs’ challenges of working with COVID-19 patients in six Viennese hospitals. The data collection took place between June 2020 and January 2021. We contacted HCWs from five public and one private hospital in Vienna – either directly or by personal introduction of the chief physician of the concerning wards (work units are detailed in table 2). In one hospital, we had a key informant who arranged further contacts to other hospital staff. The aim was to gain a maximum variation in contacts, including all kinds of HCWs (qualified nurses, nurse assistants, cleaning staff, physiotherapists, and medical doctors) in the hospital system. All other contacts were recruited by snowball sampling.

Participants were either interviewed via telephone, Webex or in person (carried out under precautionary measures of wearing a facemask and keeping distance of two meters). Those who agreed to take part in the study signed the participant consent form. All interviews were audio-recorded, except for one who felt more comfortable not being recorded. In this case, notes were taken which were subsequently sent to the participant to validate and clarify further questions. Semi-structured interviews lasted between 30 and 60 minutes and were guided by a topic guide. However, questions were adapted to the flow of the conversation and the importance the interviewees gave to a specific topic.

All transcripts were anonymized, names and any personal identifiers were removed, and interviews were labeled using a numerical code. The study was approved by the Ethics Committee of the Medical University of Vienna and the Ethics Committee of the Town of Vienna.

Transcripts were imported into Atlas.ti (Version 8.4.4) and analyzed with content analysis, using inductive and deductive coding. The deductive codes were informed by the topic guide questions; all other codes derived inductively through repeated examination of the data. Codes were united to overall themes which include lack of preparedness, physical protection, overworked personnel, staff shortage and floating staff, physical and mental effects of working in PPE, stigma, avoidance behavior and lack of recognition. The research considers changes over time with e.g. PPE shortages being more important in the beginning and overworked personnel in the current phase of the pandemic.

## 3. Results

### 3.1. Description of participants

We collected data from 30 participants who had (mainly direct but also indirect) contact with patients infected with COVID-19. Thirteen medical doctors, eleven qualified nurses and six other professions were included (table 1). 28 HCWs worked in public hospitals and two in a private hospital. Female participants predominated overall (21 female versus 9 male).

**Table 1.**
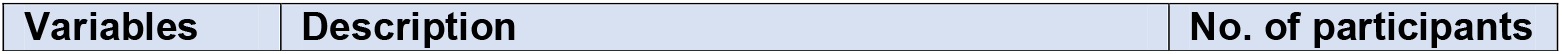
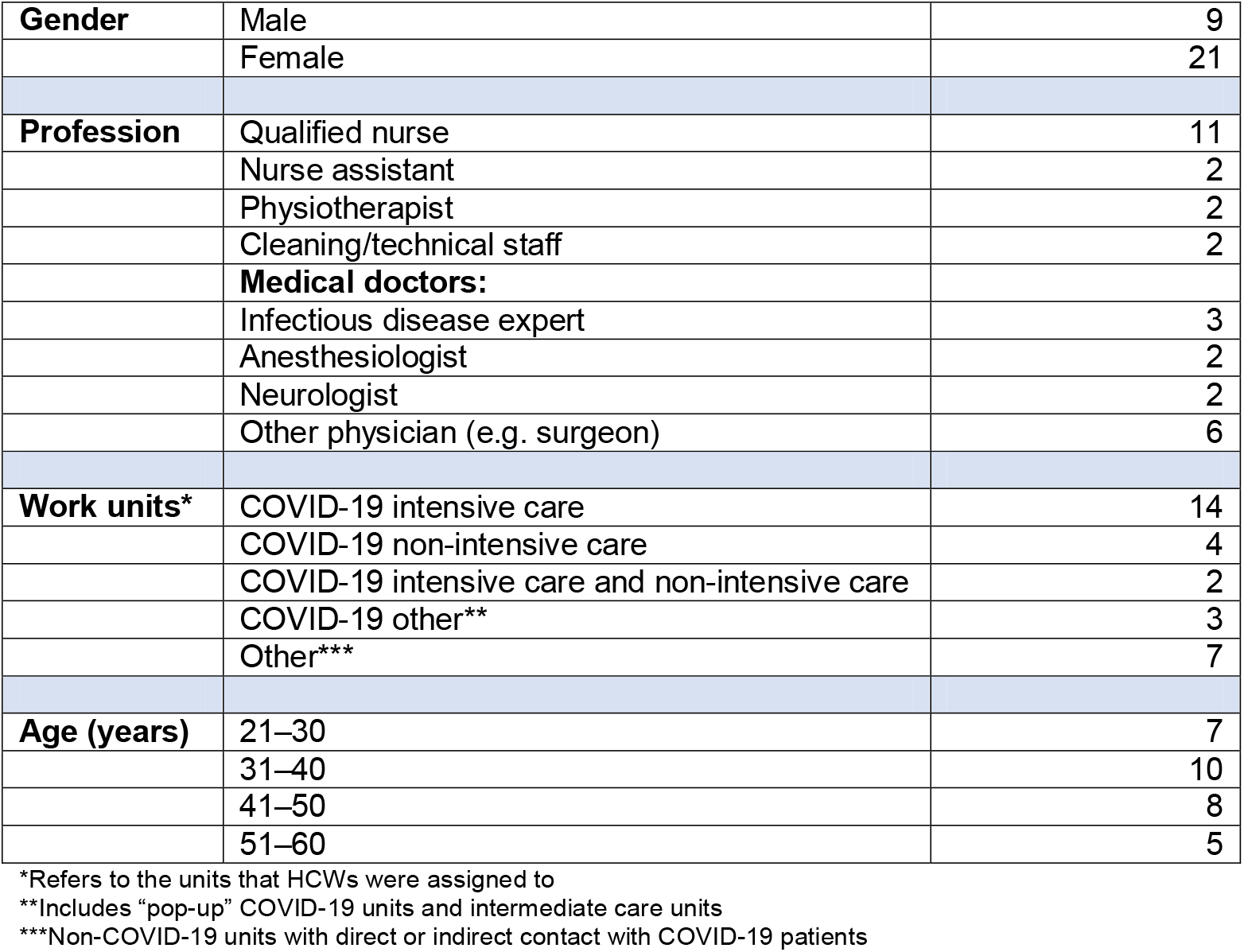
Characteristics of participants

| Variables | Description | No. of participants |
| --- | --- | --- |
| **Gender** | Male | 9 |
|  | Female | 21 |
| **Profession** | Qualified nurse | 11 |
|  | Nurse assistant | 2 |
|  | Physiotherapist | 2 |
|  | Cleaning/technical staff | 2 |
|  | **Medical doctors:** |  |
|  | Infectious disease expert | 3 |
|  | Anesthesiologist | 2 |
|  | Neurologist | 2 |
|  | Other physician (e.g. surgeon) | 6 |
| **Work units\*** | COVID-19 intensive care | 14 |
|  | COVID-19 non-intensive care | 4 |
|  | COVID-19 intensive care and non-intensive care | 2 |
|  | COVID-19 other** | 3 |
|  | Other*** | 7 |
| **Age (years)** | 21–30 | 7 |
|  | 31–40 | 10 |
|  | 41–50 | 8 |
|  | 51–60 | 5 |

\*Refers to the units that HCWs were assigned to
\*\*Includes “pop-up” COVID-19 units and intermediate care units
\*\*\*Non-COVID-19 units with direct or indirect contact with COVID-19 patients

HCWs mentioned lack of preparedness as well as the physical, mental and emotional aspects of working in this pandemic as the most challenging factors. “We had no experience; we did not know this virus. In the beginning there was no clear structure, no guidelines, no information.” (female qualified nurse 16) This statement outlines the essence of the challenges HCWs were facing while confronted with a hitherto unknown pandemic and the consequences of being ill-prepared. In addition to clinical challenges and its physical, mental and emotional implications HCWs were also troubled by delayed or unavailable infection control guidelines, uncertainties regarding physical protection, extended working hours, staff stretched to the limit and lack of supervision.

### 3.2. Lack of preparedness

HCWs referred to unavailable or delayed IPC guidelines adapted to a major infectious disease outbreak. This included for example guidelines for proper donning and doffing of PPE, guidelines for medical procedures that produce aerosols or, IPC-strategies for patient transfers. Consequently, HCWs faced many uncertainties and those units in charge of treating COVID-19 patients often had to make their own autonomous decisions.

> “How are we supposed to reanimate in this massive gear? Are we even allowed to enter with the defibrillator, as it is then contaminated (…) These were situations where we had to come up with our own guidelines on how to handle what we were faced with.” (female qualified nurse 17)
>
> “The department of hygiene gave instructions only after we became a COVID-19 ward, on the same day or maybe two days before, whilst we were already wondering about it for weeks. Examples include which respirator tools to use or the need of different filters, those which would last longer, as we don’t want to disconnect the ventilators daily because of the need to change the filter. (…) Those points were addressed super delayed by the department of hygiene.” (female qualified nurse 4)

Most participants stated that their team and their immediate superiors dealt with the situation excellently. However, complaints were made about chaos that arose on higher levels of the hospital hierarchy; participants wished for better guidance from leadership and management:

> “It is something you expect to be handled by management and not from the personnel on the ward. (…) You expect the hospital pays attention that the work procedures are properly adjusted – not that the employees take care of them.” (female medical doctor 1)

Another demanding factor was lack of coordination between hospitals. Especially in the first phase there was a lack of clarity on ICU bed capacities and the second phase during summer was considered as a missed chance to be better prepared for the predicted peak phase beginning in autumn. HCWs felt that other hospitals in Vienna were unprepared to take on COVID-19 patients. One medical doctor concluded:

> “Well, you do wonder how slow the mills in Austria are grinding and how little foresight one can have.” (male medical doctor 5)

The following statement summarizes well how little HCWs felt prepared for this demanding situation.

> “You cannot expect of staff that they have to face this situation without everything is thought through: That I am protected, I have the patients well on the monitor, I have the medicine I need or the staff that is trained in palliative care for example. (…) I do believe this could have been handled better, definitely.” (female medical doctor 28)

### 3.3. Physical protection

During the first phase of the pandemic there was a shortage of facemasks and the fear of insufficient facemasks. One medical doctor reflected upon how to define shortage of facemasks in Austria: “(…) once you start using masks that had expired 14 years ago, which we did, I would say we ran out of masks.” (male medical doctor 5) Others mentioned having used facemasks of insufficient quality or ones that did not properly fit, or that they had to reuse disposable masks. Especially FFP3 masks were rare in the beginning and often only available for rooms with patients on non-invasive ventilators where circulation of aerosols was highest.

Fear of mask shortages also led to employees stockpiling masks or that management was restrictive about their distribution: sometimes cleaning personnel or other medical support personnel were denied adequate facemasks in one hospital. One medical doctor recounted the following:

> “I have always received it (protective gear), but there was a situation that sticks well in my memory when the ward management gave me a FFP3- mask, her (the manager’s) team got a FFP3-mask because there was a suspected case, and the colleague in charge of patient transportation did not get a FFP3-mask. He got a FFP2 mask as he is only pushing the bed was the statement.” (male medical doctor 5)

Interestingly, when asked directly if they felt safe at the workplace most HCWs stated they felt sufficiently protected. Especially those working at ICUs felt better protected than in wards “outside” because they knew the infectious status of their patients and worked in protective gear. Nevertheless, most HCWs articulated fear of infecting family members.

These contradictory statements are due to different experiences among the work groups – those working in an ICU had less difficulties accessing protective equipment than cleaning or technical staff (although they also had to reuse masks). The fear of infecting family members, however, cannot be traced to these differences but may be because HCWs would accept their own infection but not an infection of their loved ones.

In addition to shortage of physical protection, there was a quest for supervision, which was often not sufficiently offered by employers. Though not everyone stated to need supportive supervision, most considered it important to be provided.

> “Of course, there are some things I miss from our employing institution as it is its responsibility to protect us. Not only to provide the protective gear but also mental protection. (…) It has the responsibility to ensure we do not get harmed mentally and physically.” (female qualified nurse 23)

### 3.4. Overworked personnel, staff shortage and floating staff

Especially in those interviews conducted around November 2020, HCWs were working over the limit of their capacities. Many reported to be mentally and physically exhausted and to need longer regeneration times than normal. HCWs not only worked extra hours or worked without having proper breaks, they also functioned in a permanent alarm mode. In addition to dealing with challenges related to infection risk and changed working procedures, HCWs faced medical uncertainties and emotional challenges due to the critical condition of patients. The difference to pre-COVID-19 was the quantity of dying patients, as for example highlighted by the metaphorical sentence “patients are dying like flies” (female qualified nurse 17). Another mental burden was seeing people without underlying medical conditions or young patients die or having to witness how patients slowly died in full consciousness and in isolation. Further, dealing with the unpredictability of the disease added to being mentally overburdened.

> “I had many patients who entered the hospital fully conscious, more or less ‘shit; now I’m one of those who has COVID-19.’ They got worse; suddenly they had a mask in their face and after 10 days under the mask in the short breaks to take a sip of water or a pill they would ask ‘I don’t know, will I recover? I feel really bad.’ (…) And they died fully conscious. I had one (patient) who had a hand mirror and he kept on checking the oxygen saturation on the monitor above his bed, he kept on looking and died with that number in his eyes. It’s like drowning. Absolutely cruel.” (female medical doctor 28)

HCWs spoke about a missing work-life balance and consequences of being overworked. Some thought to be more vulnerable to getting infected with COVID-19, others mentioned physical pain because of chronic overload.

> “We are now faced with some sick leaves. It is the high adrenalin and cortisone levels of this crisis, the ongoing emergency mode that is exhausting at some point. Basically, our bodies are giving up. (…) There are people that would like to, but they are just sick now. They do not have COVID-19, but they are sick. They have digestive issues; one has ongoing diarrhea for three weeks and he looks pale as a linen sheet.” (female medical doctor 27)

Being overworked associated with a shortage or unavailability of staff was mentioned as a main problem by study participants. Especially in autumn 2020 HCWs stated that more personnel got sick which again led to the problem of not having enough workforce at place. Strikingly, often it needed multiple requests to the administration to getting more staff.

Another related aspect was the floating of staff. To remedy staff shortages in one ward, especially qualified nurses were often recruited from other wards. Those who had free choice and switched on a voluntary basis with the option to switch back, viewed this more positively. However, many suddenly had to work with COVID-19 patients without having a professional background in infectious diseases and thus missed technical knowledge in this regard. Often, qualified nurses found themselves in a new team and there was little time to get proper training. This produced extra stress. Though also medical doctors from other units – e.g. rheumatologists – were redeployed to COVID-19 units, their situation was perceived as more stable because they could stay as a team at their unit. However, also these professions experienced a sudden shift to providing care regardless of their actual professional background.

### 3.5. Physical and mental effects of working in PPE

A recurring theme was the major physical challenge of working in protective gear. Most HCWs were not used to working in PPE. Donning and doffing of protective gear was described as time consuming and demanded high concentration to avoid any mistakes. In addition, donning and doffing was a mental burden due to the perpetual challenge of a possible infection. For example, once the facemask was sliding down in a patient room, HCWs noticed it but could not adjust the mask because they were already contaminated. This implied a high necessity of being properly prepared and adjusted before entering the patient room to work safely for two to three hours. Moreover, working in protective gear was much more exhausting. Interview partners reported heavy sweating, headaches due to wearing the equipment over longer periods of time and difficulties breathing while performing day to day clinical tasks.

Most interviewees specified that the level of care for COVID-19 patients was very high and the combination of frail patients and working with protective gear was fatiguing. One female physiotherapist (22) described that (recently) sedated patients had to be moved with all physical strengths. Protective gear made mobilizing patients, performing clinical tasks as well as communicating with patients and colleagues much more difficult.

Especially those HCWs with extensive patient contact (e.g. qualified nurses or physiotherapists) were suffering the consequences of wearing PPE for long periods of time: it drained peoples’ strengths and limited the ability to manage all patients scheduled for the day.

> "Of course, you could say, now I am leaving the room. I cannot handle it anymore. If I stay here any longer you can also admit me as a patient because I’ll collapse. But you always think: we continue and then there will be a break (…) I think the longest time in the room was 3 hours (…).” (female qualified nurse 17)

Due to self-protection measures but also to minimize exposure, the engagement with patients was limited to a minimum, and clinical tasks were bundled to use time efficiently, once in the room. This implied to constantly think ahead. Further, patient rooms could not just be entered without precautions, which put HCWs in moral distress to be unable to help patients on time in emergency situations.

### 3.6. Stigma and avoidance

HCWs experienced stigma in their private lives and observed avoidance behavior in some colleagues. Especially in the beginning of the pandemic some physicians neglected to attend patients due to the fear of getting infected.

> “We had a patient who was a cardiology patient suffering a heart attack and the cardiologists did not want to attend to the patient because they were too scared of COVID-19. You end up thinking, this is your patient who happens to have COVID-19 but it is simply not adequate patient care. Because you are scared of this stupid virus. And I keep on going in every day.” (female medical doctor 18)

One qualified nurse from another hospital did not see the nursing officer during the first months of COVID-19 and thought that her ward was being avoided. Other problems being stated were getting appointments for Computer Tomography, having X-rays done on COVID- 19 patients or getting blood examined at the laboratory. The situation improved over time mainly by constantly communicating with the concerning professions and units.

HCWs were often perceived as high-risk contacts and faced stigma in their social surroundings. Stigmatization also extended to their family members and relatives – labels such as “Coronalady” or “Corona children” give an impression of how HCWs or their family members were sometimes perceived by their social environment. Others reported that their children were not invited to friends, or personal appointments at a doctor’s office were cancelled rudely.

The predominant fear of many HCWs was to infect family members. This fear sometimes led to self-stigmatization or avoidance behavior such as sleeping in separate bedrooms, not kissing the partner or not meeting people in general. One HCW recounted that she considered herself as a role model. On the one hand, this had to do with the perception that as a HCW she should know about infection pathways. On the other hand, she directly saw the worst consequences of a COVID-19 infection. Thus, she thought that HCWs had to be especially cautious about their behavior.

> “We quickly ended up being considered contagious as well, but we are tested. Well, I think we are constantly thinking if we are imposing a risk for other people. We have all limited our social contacts to a minimum because we are afraid that we are the ones who will infect others.” (female medical doctor 26)

### 3.7. Lack of recognition

Gratitude and appreciation were important topics for most HCWs. They positively mentioned support by direct supervisors and mutual support between team members as encouraging. However, many participants missed recognition for their work by superiors at higher management levels or financial rewards (which was promised by politicians).

> "You just don’t feel valued (…) It does not have to be a monetary reward, though that would be something, because it was much more exhausting, but frankly a ‘thank you’ for showing up or saying ‘I know it is exhausting’. That is something that would qualify a leader.” (female qualified nurse 10)

One female qualified nurse (17) thought it was important that her colleagues (e.g. cleaning staff and nurse assistants) got financially remunerated as well. All of them were taking on additional or new tasks and were involved in mastering this exceptional situation.

There was a general resonance, especially among nurses, that their work (which is demanding, much needed and challenging) was not recognized by the public. They feared that their services (for the public) will fall into oblivion once the crisis is over. Further, for most HCWs appreciation by the public was largely missing. Many interview participants thought that ‘clapping at 6 pm’ did not show real appreciation or acknowledgement. HCWs stated that personalized appreciation was showing more genuine support. Take for example a banner in front of the hospital from an Austrian football club, thank you postcards or gift baskets from former patients and food delivery from restaurants.

## 4. Discussion

This study deals with occupational challenges of HCWs working during the COVID-19 pandemic. Our paper is one of only few studies dealing with this topic in the European Union. By using a qualitative exploratory approach, we outlined context-specific challenges of HCWs of different work groups. The research considered changes over time by collecting data during a period of six months, and therefore includes topics of relevance at the very beginning of the crisis and as the pandemic unfolded.

Stress factors result from structural conditions and a lack of pandemic planning on governmental/institutional level as well as clinical challenges and their physical, mental and emotional implications. Missing recognition and social stigma on a public level add to these stressors.

To put it more precisely, lack of preparedness, lack of physical protection, overworked personnel, staff shortage and reallocation of staff, physical and mental effects of working in PPE, stigma, avoidance behavior and lack of recognition were major stressors among our study participants in Vienna. These findings largely correspond with results from other international studies on related topics, showing how most experiences are shared on a global level.

In the beginning of the pandemic, lack of preparedness played a major role mainly in terms of PPE shortages and delayed IPC guidelines. PPE shortage was a global phenomenon, and the usage of inadequate PPE was also addressed in other research.^16-19^ According to a study on HCWs motivation to delivering care during COVID-19, feeling protected by the government/hospital was related to lower hesitation to work. The authors conclude that more efforts should be made on governmental/institutional level to protect HCWs, especially when it comes to preventing infections in HCWs.^20^ Another study mentions little confidence in knowledge of IPC as the main barrier to willingness to work in infectious disease outbreaks.^21^ Consequently, providing proper IPC training and adequate PPE is not only indispensable for providing a safe workplace but also influences workforce availability in the long run.

Relatedly, working in PPE entails major physical and mental strains. Long duration of wearing PPE combined with a high level of care for patients infected with COVID-19 drained HCWs’ strengths to perform their tasks. This was especially the case for qualified nurses and physiotherapists. Occupational skin injuries, difficulties breathing, or headaches were some of the physical effects of working in PPE also mentioned in other studies.^16, 19, 22^ One study outlines that HCWs continued to work despite of pain and without taking breaks. Breaks were associated with wasting scarce PPE resources and with feeling guilty, especially when there were staff shortages.^16^ This increases the need for more hospital personnel and readjustments of staff schedules to shorter shifts to ensure a safe work place.^18, 19^

In our study, staff shortage and overworked staff became routine in affecting working conditions as the pandemic unfolded. Depletion of staff affects HCWs’ mental and physical health and carries implications on workplace safety in the long run. Further, overworked personnel affect quality of care. Staff shortages and overworked staff have also been mentioned in similar research from other countries focusing on well-being of HCWs.^23-25^ One study concludes that mental health of HCWs in emergencies should be addressed with a holistic approach and a socio-ecological understanding of well-being.^24^ Multiple factors influence well-being of HCWs besides clinical challenges, for example staff shortages, taking enough rest, access to protective equipment, implementation of infection control guidelines and other external factors such as public support.^24-26^ In addition, providing contextualized psychological services is important. Interventions should be adjusted to HCWs’ specific needs such as adequate rest and availability of sufficient protective equipment.^27^ Psychological interventions should also be adapted according to sociodemographic disparities and differences among work groups.^21^ Other studies found that HCWs experience moral injury as a consequence of their commitment.^28, 29^ Insufficient protection, as well as other factors such as (lack of) actions that violate one’s ethical principles (e.g. not being able to provide good quality care to patients due to being overworked), lead to negative self-perception and distrust in the system.^28^ Once the crisis is over, a major task should therefore be after care, addressing moral injury in HCWs and rebuilding trust in the system.^29, 30^

We also found that some nurses who have no professional infectious disease background or training were recruited from other wards to remedy staff shortages. Redeployment of staff without specific training may lead to absenteeism, especially once the crisis is over.^31^ Therefore, it is even more important that redeployment is based on a voluntary decision.^26^ To tackle the problem of nurses with diverse backgrounds, experiences and skills, a Chinese hospital implemented standardized nursing procedures for work routines and work content. Other hospitals implemented clearly defined responsibilities of staff and training programs for protective measures and handling equipment.^18, 32^

Social stigma and self-stigmatization mainly occurred outside the hospital but added to the mentioned stress factors. COVID-19 related stigmatization and attacks on HCWs are a global social consequence of this pandemic. In many countries stigmatization of HCWs and experiences of being avoided or insulted posed a threat to HCWs and often resulted in violence and harassment.^33-35^ This is especially worrisome as HCWs respond to a health crisis to save lives and protect the society while exposing themselves to the risk of infection.^36^ Stigma and self-stigmatization may be exacerbated by the questions of guilt - who is responsible for another person’s infection or death - which seems to be a characteristic of this pandemic. The governmental measures of social distancing, which are necessary to diminish infection rates, make boundaries between social distance and social stigma less tangible.

Another sensitive and related topic is avoidance of colleagues to treat COVID-19 patients, which was relevant in our interviews at the beginning of the pandemic. To our knowledge, this finding has not been addressed by other authors up to date. This avoidance may stem from anxiety to infect oneself or family members, from respect of this formerly unknown threat and limited scientific knowledge available. It may also be the result of not feeling properly prepared to work in an infectious disease context. While some HCWs are more resilient to working in this exceptional situation, it may be harder and more burdensome for others. Avoidance behavior of colleagues needs to be addressed as it may lead to conflicts, additional workload, undertreatment and delayed patient care of already vulnerable patients.

Finally, HCWs felt a lack of recognition, acknowledgement and appreciation for working under such very specific circumstances. This includes financial compensation but also immaterial rewards such as showing gratitude from higher management levels and personalized appreciation by politicians and the public. Showing gratitude to HCWs and acknowledging their working conditions is one of the key elements of protecting mental health of HCWs and is known as fostering resilience.^30, 37^

### Limitations

We focused on the core topics but are aware that stressors of HCWs are more complex. Doing research during a pandemic posed several challenges, including reaching gatekeepers and interview partners. Interviews sometimes took place under rushed conditions, after clinical work and with overworked or tired HCWs. Consequently, some interviews may have been shorter than usual or may not have yielded in-depth considerations. Further, there might be more differences in experiences between professions and different occupational groups which we were not able to consider.

## 5. Conclusions and recommendations

Despite the medical difficulties and unpredictable aspects of a pandemic can hardly be prepared for, it is necessary to ensure a structural framework, e.g. with guidelines and standard operating procedure, in order for HCWs to feel prepared, protected and cared for. This framework is also needed to ensure optimized psycho-social working conditions of HCWs and support in these challenging times.

In our context, mainly organizational-level recommendations are necessary to prepare for later phases of the pandemic or new emerging threats. Managing the shortages on multiple levels will be paramount. Four themes are of importance: Firstly, to tackle the shortage of PPE and therefore ensure physical protection. Secondly, to mitigate shortage of human workforce and averting chronic occupational overload. Adequate providing of medical personnel, especially nursing staff, is essential. Voluntarism plays an important role in terms of redeployment of staff and HCWs should be given the option to switch back or at least take personal preferences into account. Thirdly, proper training and education in IPC (including timely providing of necessary guidelines) but also palliative care are important. Cleaning and service staff should receive tailored IPC training and education to cope with fears and to be safe. In general, professionally handling and addressing fear is needed to overcome avoidance behavior. Simulation exercises for both doctors and qualified nurses and professional debriefing could better prepare HCWs for stressful situations. Lastly, caring for HCWs mental health is essential, especially offering supportive supervision convenient to HCWs’ working schedule. Gratitude from superiors, politicians and the public are indispensable for showing support and foster resilience.

## Data Availability

Due to the nature of this research, participants of this study did not agree for their data to be shared publicly, so supporting data is not available.

